# National Burden of Disability in Bangladesh and its Socio-Demographic Correlates

**DOI:** 10.1101/2023.09.13.23295500

**Authors:** Mizanur Rahman, Md Shohel Rana, Gulam Khandaker, Md Mostafizur Rahman, Md Nuruzzaman Khan

## Abstract

**Background:** The burden of disability remains a global challenge, particularly in low- and middle-income countries (LMICs) like Bangladesh. Understanding the national-level burden of disability and its socio-demographic determinants is crucial for informed policy and program development. We aims to explore the national level burden of disability in Bangladesh and its socio-demographic correlates.

**Methods:** This study presents a cross-sectional analysis of 155,025 participants in the 2021 National Survey on Persons with Disabilities (NSPD). Disability status (yes, no) served as the outcome variable. The explanatory variables considered were factors at the individual, household, and community levels. A multilevel mixed-effect logistic regression model was used to explore the explanatory variables associated with the outcome variable, splitting the total sample into two age-based groups: 0-17 years and 18-95 years.

**Results:** Our findings reveal a disability prevalence of nearly 3% in Bangladesh. The prevalent forms of disability encompassed physical disability and visual impairment. Close to one in ten individuals with disabilities in Bangladesh reported experiencing more than one type of disability concurrently. The likelihoods of disability were observed to be higher among individuals with higher educational attainment, those belonging to households with elevated socio-economic status, those engaged in white-collar occupations, and those residing in the Khulna, Rajshahi, and Rangpur divisions. Additionally, a heightened likelihood of disability was observed among communities with moderate to higher illiteracy rates.

**Conclusion:** The implications of these findings extend extensively to policy formulation and the allocation of resources aimed at tackling the multifaceted challenges encountered by persons with disability.

**Research in Context:** *Evidence before this study:* Previous research has recognized the substantial burden of disability in Bangladesh, yet lacked comprehensive nationwide assessments of its prevalence and socio-demographic factors. Existing studies often focused on localized contexts, leaving a gap in understanding the broader landscape of disability within the country.

*Added value of this study:* This study substantially contributes to the understanding of disability in Bangladesh by conducting a comprehensive national-level analysis. Using representative samples and rigorous methodology, it explores diverse dimensions of disability and its socio-demographic factors. This comprehensive approach bridges the gap in existing literature, offering insights into the nuanced intricacies of disability prevalence and correlates.

*Implications of this study findings:* The study’s findings have far-reaching implications for policy and resource allocation. By uncovering disability prevalence and its socio-demographic associations, the study enables policymakers to formulate targeted interventions, addressing challenges across education, employment, healthcare, and social inclusion. Moreover, contributing to the global disability discourse, the study underscores the significance of context-specific investigations for impactful interventions. These insights can shape disability-related policies and programs, not just in Bangladesh but also in comparable socio-economic settings worldwide.

## Introduction

People living with disabilities (PWD) constitute the world’s largest minority group, with an estimated population of approximately 1.3 billion globally as of 2022 ^1^. Over 80% of them reside in low- and middle-income countries (LMICs) ^2^. Notably, these figures are undergoing a rapid escalation on a global scale, particularly within LMICs, driven by the increasing numbers of the aging population and rising prevalence of disabilities among them ^1,2^. Furthermore, LMICs are currently witnessing a swift surge in urbanization and industrialization, leading to a growing workforce in urban jobs and construction sectors, often lacking proper safety oversight ^3^. This situation has resulted in a surge of accidents and injuries, accompanied by a rise in fatalities ^4^. Additionally, road-traffic injuries are becoming more prevalent in LMICs, contributing to a grim statistic of 1.35 million global deaths due to road traffic crashes in 2020, with over 90% of these tragic incidents occurring within LMICs ^5,6^. Nearly 50 million additional individuals sustain non-fatal injuries, a significant proportion of whom acquire disabilities as a consequence of their injuries ^7^. Notably, the rapid advancement of medical technology is profoundly influencing the trajectory of this trend, shaping the landscape of disability and injury outcomes ^8^. Bangladesh plays a significant role in this overarching trend, positioned as the eighth most populous country globally, boasting a population nearing 168 million ^7^. The nation is undergoing rapid urbanization and industrialization, consequently witnessing the concurrent emergence of road-traffic injuries as an everyday occurrence ^3,9^. These burdens in Bangladesh are compounded by the persistently high rates of poverty and engagement in hazardous occupations, which inherently elevate the likelihood of casualties ^10^.

Individuals with disabilities necessitate heightened support for their livelihoods, particularly within LMICs, where they frequently encounter marginalization even within their own families and heavily rely on social assistance ^11–13^. The fiercely competitive job market and non-disability friendly communication and transportation system further amplifies their vulnerability and increase their dependency on social support to sustain their livelihoods ^14,15^. This underscores the pressing need to delve into the national scope of the disability burden and its intricate socio-economic interconnections. This exploration is imperative, as it equips relevant stakeholders with essential insights for informed planning and program implementation ^12,13^. However, such comprehensive assessments are often lacking at the country level, and the global evidence frequently falls short in encapsulating the true prevalence and nuances of disability dynamics ^1,16^. Furthermore, the available evidence at the country level frequently generates contradictory results due to disparate approaches in measuring disability, limited sample sizes, and imprecise methodologies ^12,17^. For instance, in Bangladesh, to our knowledge, no study has comprehensively investigated the nationwide disability burden and its associated correlates. Existing studies have reported widely conflicting figures for the burden of disability (ranging from 1.0% to 18.0%) ^18,19^. Coupled with the fact that disability experiences are often uniquely country-specific and could not depend on other nations’ estimates ^20^, this inconsistency poses a formidable challenge to policymakers and program implementers in Bangladesh. To bridge this significant gap, our study aims to precisely examine the national-level disability burden and its socio-demographic correlates, offering a more accurate and contextually relevant understanding of the multifaceted dimensions of people with disability in Bangladesh.

## Methods

### Sampling strategy

Data were obtained from the 2021 National Survey on Persons with Disabilities (NSPD) conducted by the Bangladesh Bureau of Statistics (BBS). This survey employed a two-stage stratified random sampling technique to identify the participants. In the initial stage, 800 primary sampling units (PSUs) were selected at random from the 2011 Bangladesh National Population Census’s list of 293,579 PSUs. Subsequently, 45 households were systematically chosen from each of the initially selected PSUs in the second sampling stage. This approach yielded a roster of 36,000 households, with data collected from 35,493 households, attaining a robust coverage rate of 99.9%. The survey encompassed all usual residents of the chosen households, amounting to 14,659 children aged 0-4 years, 39,513 children aged 5-17 years, and 100,853 adults aged 18-95 years, totaling 155,025 participants. All were included in our analysis. For a comprehensive understanding of the survey’s sampling procedure, further information is available in the respective survey report ^21^.

### Outcome variables

The outcome variable under consideration was the respondents’ disability status. During the survey, participants were inquired about the presence of any form of disability. They were presented with a predefined list of options to choose from. If the disability they experienced was not listed, they were given the opportunity to select “other” and write the form of disability they are experiencing. The provided list encompassed disabilities such as autism, physical disability, mental illness disability, visual impairment, speech impairment, intellectual disability, hearing impairment, hearing-visual impairment, cerebral palsy, down syndrome, multidimensional disability, and other. These options were developed based on the framework of disability rights protection established in Bangladesh in 2003 and the Washington Group on Disability Statistics Model ^22^. We reclassified the responses into two categories: individuals with disabilities (coded as 1) if respondents indicated at least one type of disability, and individuals without disabilities (coded as 0) if respondents reported the absence of any disability.

### Explanatory variables

We embarked on a comprehensive two-stage selection process to incorporate an extensive array of explanatory variables. Initially, we compiled a list of potential explanatory variables derived from an exhaustive review of existing literature ^15,23–25^. Subsequently, these identified variables were cross-referenced with the survey data to verify their availability. Those variables that were indeed present were subsequently classified according to the socio-ecological model of health into three tiers: individual, household, and community-level factors. The individual-level factors encompassed the respondent’s age (0-17 years, 18-59 years, and ≥60 years), educational attainment (no education, primary, secondary, and higher), gender (male or female), occupation (agriculture, blue-collar work, pink-collar work, white-collar work, student, housewife, and others), and marital status (married, unmarried, or widowed/divorced/separated). Additionally, at the household level, a variable representing wealth quintiles was included. This variable was formulated by the survey authority via principal component analysis of household asset-related variables within the respondent’s residing household, such as roofing type and ownership of a refrigerator. Moving to community-level factors, we integrated the respondents’ place of residence (urban and rural) and their division (Barishal, Chattogram, Dhaka, Khulna, Mymensingh, Rajshahi, Rangpur, and Sylhet). Two other community-level variables considered were community-level illiteracy and community-level poverty. The data for these variables were not directly available in the survey. However, we calculated them by using respondents’ responses to community-level illiteracy and community-level poverty at the PSU level. The comprehensive details regarding the procedure for creating these variables have been documented in a separate publication ^26^.

### Statistical analysis

Descriptive statistics were used to provide an overview of both the respondents’ characteristics and the presence of disabilities. The prevalence of disability was calculated by dividing the total number of individuals with disabilities in a particular group or overall by the total population. To examine the variation in disability status across the considered explanatory variables and assess their statistical significance, cross-tabulations were executed. The determination of statistical significance was carried out using Chi-square tests. For delving into the associations between the outcome variables and explanatory variables, multilevel mixed-effect logistic regression model was used. The rationale behind selecting the multilevel modeling approach is the nested structure of the NSPD data. This data configuration entails individuals being nested within households, which in turn are nested within PSUs. Prior research has indicated that multilevel modeling yields more robust outcomes when dealing with clustered data of this nature, in comparison to conventional simple logistic regression models ^27^. In the process of model development, the total sample was stratified into two categories: ages 0-17 and ages 18 and above. For each of these age groups, a progressive model building technique was employed, involving the creation of four distinct models: null model, individual-level model, household-level model, and full model. The null model included only the disability status whereas in the individuals level model, respondents’ characteristics were considered. Households level characteristics were incorporated at the household level model along with individual level factors. The final model included all individual, households and community level factors. The results are reported as Odds Ratios, accompanied by their corresponding 95% Confidence Intervals (95% CI). The statistical analyses were carried out using STATA/SE 14.0 (Stata Corp LP, College Station, Texas, United States of America).

### Ethics approval

The survey we analyzed was reviewed and approved by the Ethics Committee of the Bangladesh Bureau of Statistics. We obtained de-identified data from the BBS through an existing research proposal. Therefore, there is no need for any additional ethical approval.

## Result

### Background characteristics of the respondents

The demographic characteristics of the respondents are presented in Table 1. Out of the 155,025 individuals whose data were analyzed, 4,293 individuals exhibited some form of disability, constituting approximately 2.79% of the overall sample. The average age of the respondents was 28.89, with nearly one-third of the total respondents being 17 years old or younger. Just over one-third of the entire respondent population lacked formal education, and roughly 26% identified as students. Approximately a quarter of all individuals were categorized as housewives. A significant majority, accounting for over 78% of the total respondents, reported residing in rural areas, while around 24% indicated Dhaka as their region of residence.

**Table 1:**
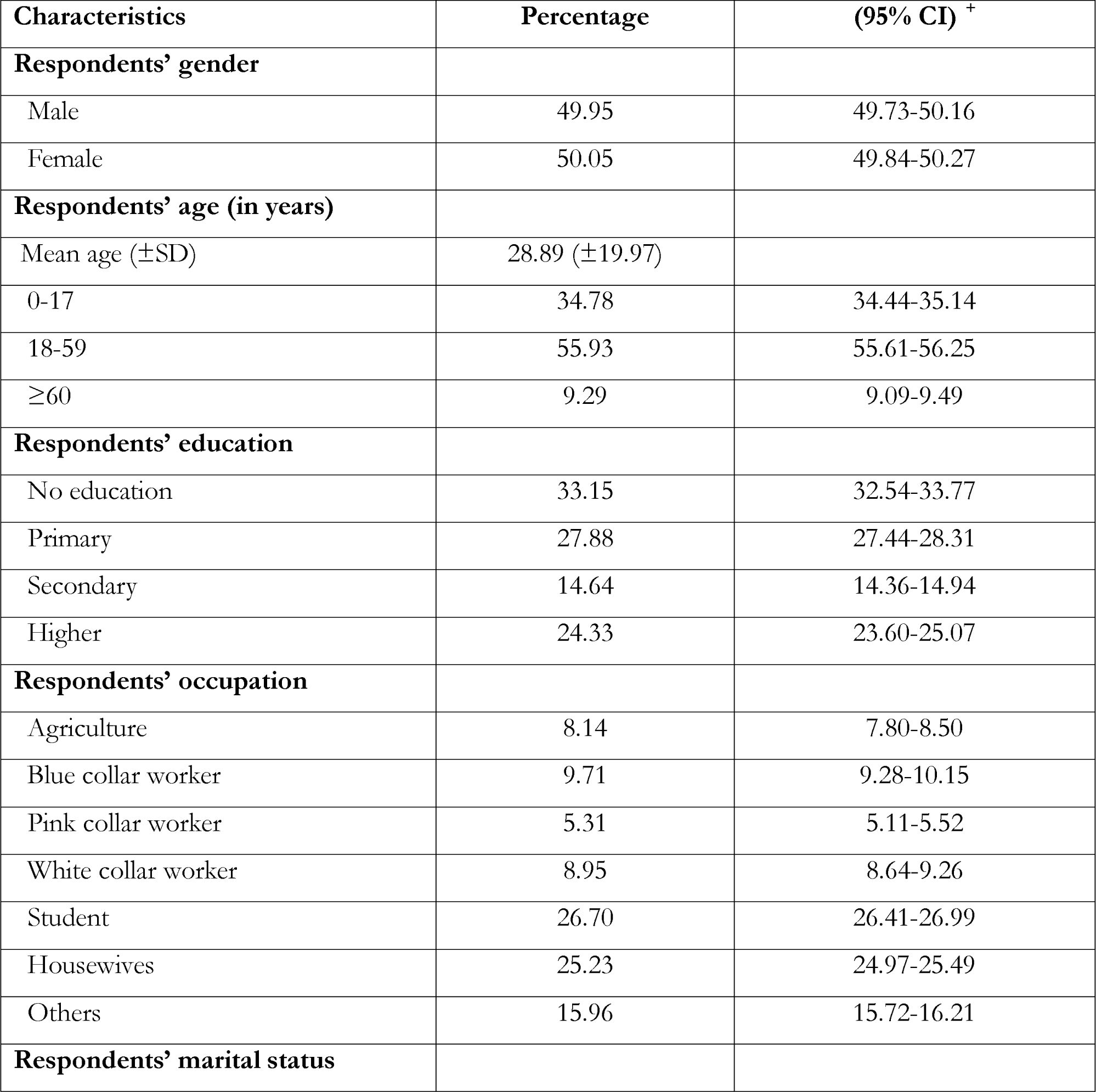
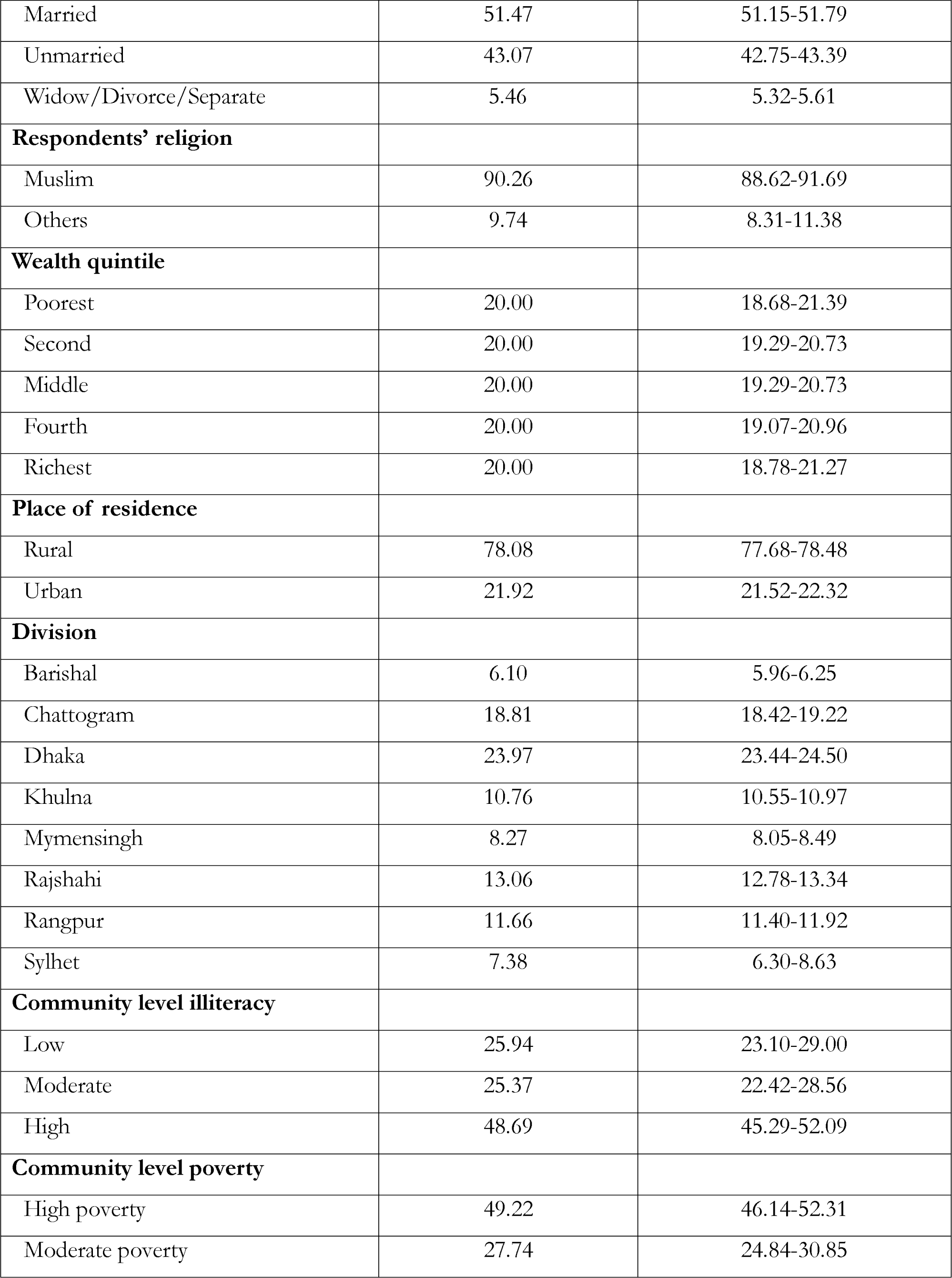
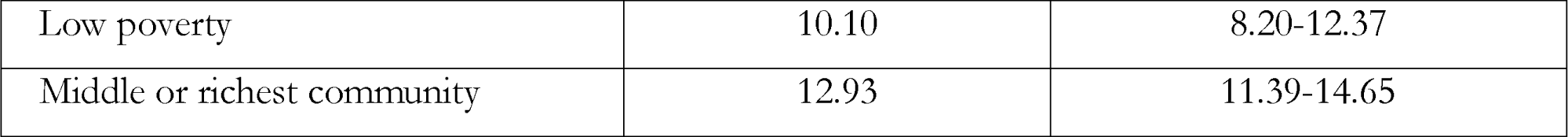
Background characteristics of the study population, N=155,025, Bangladesh, 2021.

### Prevalence of different types of disability in Bangladesh

Table 2 presents the distribution of individuals with disabilities according to the type of disability. The most prevalent form of disability was physical disability, which was reported by nearly 42% of the total respondents. Other notable categories of disabilities included visual impairment (14.11%). Nearly 12% of the total PWD reported more than one form of disability concurrently.

**Table 2:** Type of impairments in disability domain”, Bangladesh, N=4,293, Bangladesh.

| **Disability types** | **Prevalence, 95% CI** |
| --- | --- |
| Autism or autism spectrum disorder | 1.60 (1.25-2.05) |
| Physical impairment | 42.44 (40.79-44.11) |
| Mental illness | 8.38 (7.54-9.30) |
| Visual impairment | 14.11 (13.03-15.25) |
| Speech impairment | 4.08 (3.48-4.77) |
| Intellectual disability | 4.92 (4.25-5.69) |
| Hearing impairment | 6.97 (6.15-7.90) |
| Hearing-visual impairment | 0.46 (0.30-0.71) |
| Cerebral palsy | 2.77 (2.31-3.32) |
| Down syndrome | 1.22 (0.91-1.62) |
| Multidimensional disability | 11.60 (10.55-12.75) |
| Others | 1.45 (1.06-1.96) |

### Prevalence of disability and individual, household and community level factors

The presentation of disability prevalence across the considered explanatory variables are presented in Table 3. A higher prevalence of disability was found among males, individuals aged 60 or older, those without formal education, and those classified as widowed, divorced, or separated. The most substantial prevalence of disability was found in the Khulna and Rangpur divisions. Our further exploration on the prevalence of disability at the more nuance level, district level (second administrative level in Bangladesh) presents evidence of district level variations in disabilit prevalence (Figure 1). Significant variations in the occurrence of disability was found individual, household, and community-level factors considered as explanatory variables except respondents’ religion and community level illiteracy.

**Table 3:** Distribution of disability and individual, household and community level factors, Bangladesh.

| Characteristics | Persons with disability |  | p-value |
| --- | --- | --- | --- |
|  | Yes | No |  |
| **Respondents' gender** |  |  |  |
| Male | 3.28 | 96.73 | p<0.000 |
| Female | 2.32 | 97.69 |  |
| **Respondents' age (in years)** |  |  |  |
| 0-17 | 1.67 | 98.33 | p<0.000 |
| 18-59 | 2.55 | 97.45 |  |
| ≥60 | 8.49 | 91.51 |  |
| **Respondents' education** |  |  |  |
| No education | 4.73 | 95.27 | p<0.000 |
| Primary | 2.40 | 97.60 |  |
| Secondary | 1.62 | 98.38 |  |
| Higher | 1.32 | 98.68 |  |
| **Respondents' occupation** |  |  |  |
| Agriculture | 3.29 | 96.71 | p<0.000 |
| Blue collar worker | 1.95 | 98.05 |  |
| Pink collar worker | 2.01 | 97.99 |  |
| White collar worker | 2.73 | 97.27 |  |
| Student | 1.18 | 98.82 |  |
| Housewives | 1.30 | 98.70 |  |
| Others | 8.41 | 91.59 |  |
| **Respondents' marital status** |  |  |  |
| Married | 2.61 | 97.39 | p<0.000 |
| Unmarried | 2.28 | 97.73 |  |
| Widow/Divorce/Separate | 8.66 | 91.34 |  |
| **Respondents' religion** |  |  |  |
| Muslim | 2.77 | 97.23 | p=0.139 |
| Others | 3.01 | 96.99 |  |
| **Wealth quintile** |  |  |  |
| Poorest | 3.79 | 96.21 | p<0.000 |
| Second | 3.07 | 96.93 |  |
| Middle | 2.78 | 97.23 |  |
| Fourth | 2.37 | 97.63 |  |
| Richest | 1.98 | 98.02 |  |
| **Place of residence** |  |  |  |
| Rural | 2.89 | 97.11 | p<0.000 |
| Urban | 2.44 | 97.56 |  |
| **Place of region** |  |  |  |
| Barishal | 2.42 | 97.58 | p<0.000 |
| Chattogram | 2.41 | 97.59 |  |
| Dhaka | 2.51 | 97.49 |  |
| Khulna | 3.61 | 96.39 |  |
| Mymensingh | 2.45 | 97.55 |  |
| Rajshahi | 3.30 | 96.7 |  |
| Rangpur | 3.54 | 96.46 |  |
| Sylhet | 2.15 | 97.85 |  |
| **Community level illiteracy** |  |  |  |
| Low | 2.73 | 97.27 | p=0.3002 |
| Moderate | 2.95 | 97.05 |  |
| High | 2.75 | 97.25 |  |
| **Community level poverty** |  |  |  |
| High poverty | 2.97 | 97.03 | p<0.000 |
| Moderate poverty | 2.84 | 97.16 |  |
| Low poverty | 2.65 | 97.35 |  |
| Middle or richest community | 2.15 | 97.85 |  |

**Figure 1:**
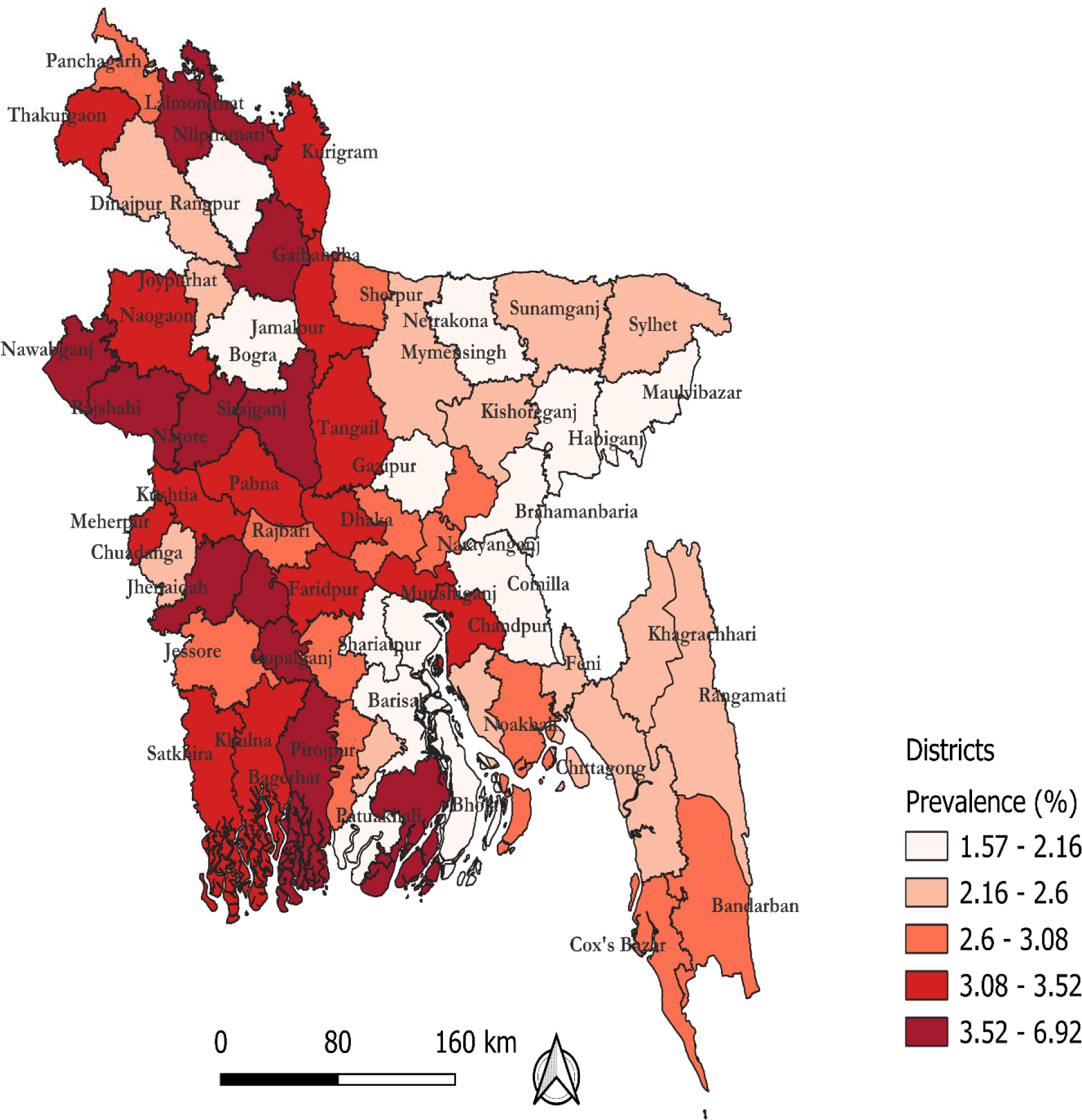
District-wise prevalence of disability in Bangladesh.

### Individuals, households, and community level characteristics associated with disability

The factors associated with disability were evaluated using a multilevel mixed-effect logistic regression model. Models were constructed for two distinct age groups: 0-17 years (first model) and 18 years or older (second model). For each sample category, a total of four models were run, with the optimal model determined through a comparison of the Intra Class Correlation (ICC), Akaike Information Criterion (AIC) and Bayesian Information Criterion (BIC). The best model was one which had the lowest ICC, AIC and BIC. The full model met these criteria, as such it was selected as the best fitted model (Table 4). The results of final models for both groups are presented in Table 5 whereas the results of full models are presented in the supplementary table 1 and 2.

**Table 3:**
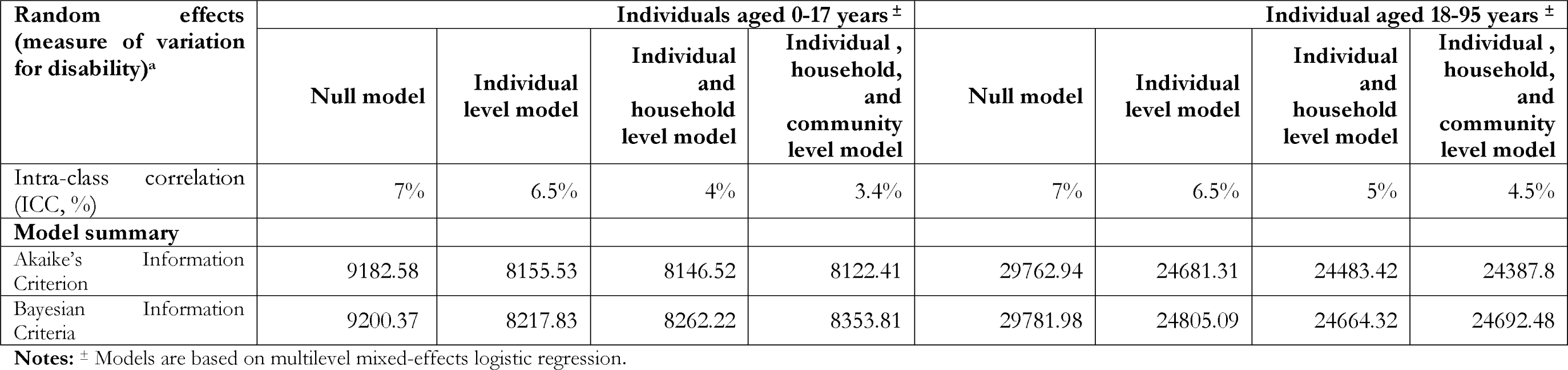
Results from the random intercept model (measure of variation) at cluster/community level for persons with disability aged 0-17 years and aged 18-95 years in Bangladeshi population, NSPD 2021.

**Table 4:** Individual, households and community level factors associated with disability in Bangladesh.

| Characteristics | Odds ratio (95% Confidence Interval) |  |
| --- | --- | --- |
|  | Individuals aged 0-17 years | Individual aged 18-95 years |
| **Respondents' age (in years)** | 0.74 (0.73-0.75) *** | 1.00 (1.00-1.00) |
| **Respondents' gender** |  |  |
| Male | 1.00 | 1.00 |
| Female | 1.00 (0.86-1.14) | 1.08 (0.96-1.21) |
| **Respondents' education** |  |  |
| No education | 0.01 (0.01-0.02) *** | 0.21 (0.19-0.25) *** |
| Primary | 0.10 (0.07-0.15) *** | 0.36 (0.32-0.42) *** |
| Secondary | 0.35 (0.23-0.52) *** | 0.54 (0.46-0.64) *** |
| Higher | 1.00 | 1.00 |
| **Respondents' occupation** |  |  |
| Agriculture | na | 1.00 |
| Blue collar worker | na | 1.42 (1.21-1.67) *** |
| Pink collar worker | na | 0.97 (0.80-1.17) |
| White collar worker | na | 0.80 (0.68-0.93) ** |
| Student | na | 1.51 (1.16-1.97) *** |
| Housewives | na | 1.72 (1.45-2.05) *** |
| Others | na | 0.12 (0.10-0.13) *** |
| **Respondents' marital status** |  |  |
| Married | 1.00 | 1.00 |
| Unmarried | 0.22 (0.09-0.51) *** | 0.42 (0.37-0.48) *** |
| Widow/Divorce/Separate | 0.23 (0.02-2.30) | 0.94 (0.83-1.06) |
| **Wealth Quintile** |  |  |
| Poorest | 1.00 | 1.00 |
| Second | 1.07(0.88-1.30) | 1.18 (1.06-1.32) *** |
| Middle | 1.22 (0.98-1.51) | 1.30 (1.16-1.45) *** |
| Fourth | 10.29 (1.01-1.64) ** | 1.49 (1.31-1.69) *** |
| Richest | 1.36 (1.01-1.82) ** | 1.65 (1.40-1.95) *** |
| **Place of residence** |  |  |
| Rural | 1.00 | 1.00 |
| Urban | 1.14 (0.87-1.50) | 0.97 (0.82-1.14) |
| **Place of region** |  |  |
| Barishal | 1.00 | 1.00 |
| Chattogram | 1.19 (0.86-1.66) | 0.92 (0.75-1.13) |
| Dhaka | 1.00 (0.71-1.40) | 0.85 (0.69-1.04) |
| Khulna | 0.68 (0.48-0.97) ** | 0.65 (0.52-0.80) *** |
| Mymensingh | 1.10 (0.76-1.57) | 0.96 (0.76-1.21) |
| Rajshahi | 0.79 (0.57-0.1.11) | 0.63 (0.51-0.77) *** |
| Rangpur | 0.64 (0.46-0.89) *** | 0.59 (0.48-0.72) *** |
| Sylhet | 1.10 (0.77-1.58) | 1.23 (0.98-1.56) |
| **Community level illiteracy** |  |  |
| Low | 1.00 | 1.00 |
| Moderate | 0.94 (0.75-1.18) | 1.23 (1.07-1.42) *** |
| High | 1.23 (0.99-1.55) | 1.40 (1.22-1.61) *** |
| **Community level poverty** |  |  |
| High poverty | 1.00 | 1.00 |
| Moderate poverty | 0.91 (0.74-1.12) | 1.06 (0.93-1.21) |
| Low poverty | 0.92 (0.65-1.30) | 1.01 (0.82-1.24) |
| Middle or richest community | 0.71 (0.47-1.07) | 1.14 (0.88-1.49) |

**Note:** \*\* p<0.05, \*\*\* p<0.01, na= Not applicable

In the 0-17 model, one-year increase in age was linked with a 26% reduction (aOR: 0.74, 95% CI: 0.73-0.75) in the likelihood of disability. Consistent trends emerged in both the first and second models with regards to respondents’ education. The likelihood of disability decreased progressively with diminishing levels of education, particularly in comparison to those with higher education. Contrasting occupation categories, the odds of disability were higher among blue-collar workers (aOR: 1.42, 95% CI: 1.21-1.67), students (aOR: 1.51, 95% CI: 1.16-1.97), and housewives (aOR: 1.72, 95% CI: 1.45-2.05) compared to individuals engaged in agriculture. Conversely, lower odds of disability were observed among white-collar workers (aOR: 0.80, 95% CI: 0.68-0.93) and individuals falling into the “others” category (aOR: 0.12, 95% CI: 0.10-0.13). The likelihood of disability was 78% lower (aOR: 0.22, 95% CI: 0.09-0.51) among unmarried individuals aged 0-17 years and 48% lower (aOR: 0.42, 95% CI: 0.37-0.48) among the unmarried individuals aged 18 and more as compared to their married counterparts. Moreover, the odds of disability increased with ascending household wealth quintiles, indicating a gradient effect. Individuals residing in the Rangpur, Rajshahi, and Khulna divisions demonstrated lower odds of disability compared to those in the Barishal division. Notably, residing in a community with moderate (aOR: 1.23, 95% CI: 1.07-1.42) or high (aOR: 1.40, 95% CI: 1.22-1.61) illiteracy levels was associated with a 123-140% heightened likelihood of disability when contrasted with residing in a community with lower illiteracy levels.

## Discussion

The objectives of this study were twofold: to investigate the national-level burden of disability in Bangladesh and to identify the associated factors. Our findings reveal that approximately 3% of Bangladesh’s total population experiences some form of disability. The prevalent forms of disability are physical disability, followed by visual impairment and multidimensional disability. Among the demographic segments, higher disability prevalence is observed among males, individuals aged 60 or older, those with no formal education, and those residing in the Khulna, Rajshahi, and Rangpur divisions. Conversely, lower odds of disability were found among unmarried individuals, individuals with comparatively lower educational attainment, and respondents in the Khulna and Rangpur divisions. Conversely, a higher likelihood of disability was identified among respondents with relatively improved household wealth quintiles and those living in communities with moderate to high levels of illiteracy. Notably, these findings hold considerable robustness as they emerge from an advanced statistical model applied to a large, nationally representative sample. The model accounts for a comprehensive range of individual, household, and community-level factors. Therefore, insights reported in this study are poised to inform national-level policymaking and program development.

The documented disability prevalence of approximately 3% in the present study conducted in Bangladesh starkly contrasts with the 16% average disability rate at the global level reported by the WHO in 2022 ^1^. Notably, disability prevalence exhibited substantial variation at the national level, ranging from 1.0% to 18.0% ^19^. This discrepancy highlights a remarkable advancement within the country’s context, although the absolute number of individuals grappling with disabilities remains significant, encompassing an estimated 4.8 million people ^15^. The underlying factors contributing to this relatively diminished prevalence in Bangladesh can be attributed to the comparatively smaller proportion of the aging populace and a higher concentration of rural residents ^2,24^. It is essential, however, to acknowledge the ongoing rapid shifts observed in both these indicators within the nation. The surge in urban dwellers and the aging population signals the potential for an escalation in disability prevalence in the forthcoming years ^28^. This projection can be attributed to a surge in accidents stemming from urbanization and industrialization, alongside the migration from rural to urban settings ^3,7^. Additionally, heightened survival rates subsequent to accidents leading to physical impairments, coupled with increased life expectancy amidst various health conditions, collectively contribute to this trajectory ^6,29^.

These characteristics are also exhibit a more pronounced presence among individuals with comparatively higher educational backgrounds and white-collar occupations in Bangladesh and other LMICs ^15,30^. Moreover, the gradual escalation of disability likelihoods corresponding to household wealth quintiles provides further corroboration for the aforementioned suppositions. While seemingly paradoxical, these observations can be elucidated through a nuanced lens. Increased awareness and accessibility to healthcare services among people with these characteristics can lead to heightened diagnosis rates ^31^. Moreover, heightened educational levels and enhanced socioeconomic status often correlate with increased engagement in activities involving greater risk, whether due to occupation-related hazards or participation in recreational pursuits ^30^. Paradoxically, despite having improved access to healthcare services, there is a prevailing norm that prioritize the receipt of health care services based on immediate needs, seeking medical attention solely when unwell, rather than prioritizing services that hold potential for sustained long-term health benefits ^8,31^. It allows certain conditions to evolve into disabilities over time. Moreover, the extended lifespan typically associated with enhanced educational and economic circumstances can predispose individuals to age-associated disabilities ^31^. Psychosocial stressors, a facet frequently encountered within higher socioeconomic tiers, might also contribute to mental health challenges that can, over time, manifest as disabilities ^12^.

The persistent burden of non-communicable diseases in Bangladesh, akin to that in other LMICs, is particularly pronounced among individuals with higher educational achievements, residing in urban areas, and positioned within the upper echelons of wealth quintiles ^28,32^. The primary underlying reasons are an increased dependency on Western food and a higher rate of overweight/obesity ^33^. Individuals belonging to these categories tend to adopt more hygienic lifestyles, often allocating less time to tending to their physical well-being. People with these characteristics are more likely to report disability, as indicated in this study and in other studies conducted in LMICs ^34^. A recent Bangladeshi student using the same cohort of disabled individuals we analyzed also reported that around half of them have comorbidities, with highly prevalent comorbidity being chronic conditions, including NCDs ^35^. The pathways of a higher rate of disability following chronic conditions could be linked with a higher rate of undiagnosed and untreated chronic conditions, with treatment received when disability has already onset.

However, higher likelihoods of disability among individuals in the comparatively lower education and resided in the Khulna, Rajshahi and Rangpur divisions indicate different perspective of these observations. As per the current community structure in Bangladesh, people in the community with lower education predominantly engaged in risky job, have very poor knowledge about health status and likely to accept healthcare services. Similarly, the aforementioned divisions are characterized by lower prevalence of urbanization and industrialization, with inhabitant there are primarily engaged with agriculture ^3^. They are also less likely to access healthcare services indicating risk of any health conditions are primary undiagnosed and untreated indicating increased likelihoods of disability ^31^. We found a very higher percentage of disability among widowed, divorced, or separated as compared to the normal married or unmarried women. However, the likelihood of disability among this group was found insignificant. This could be attributed to factors such as economic hardship, social isolation, depression, anxiety, and chronic health conditions ^36,37^. Unmarried women may face financial challenges, leading to stress and health issues ^37^. They may also have limited social support, contributing to stress-related health problems. The loss of a spouse or the end of a marriage can trigger depression and anxiety, potentially leading to disability ^38^. Unmarried women might also have a higher prevalence of chronic health conditions like heart disease, stroke, and diabetes, which can further increase the risk of disability ^37,39^. However, there could be other factors involved and further research is necessary to fully understand this phenomenon.

The policy implications of this study is that national actual burden of person with disability is higher with nearly 3% disability which is belong to around 4.6 million people ^21^. The likelihoods of being disabled are higher among comparatively higher educated, higher wealth quintile and better occupation holders. This indicates lower awareness regarding health status ^40^. This suggest needs for awareness building programs to accelerate healthcare services use in regular basis rather than accessing healthcare services when feel unwell.

This study has several strengths. As far we know this is the first study in Bangladesh that explored the national level burden of disability and its corelates based on quite large sample collected by a nationally representative households survey. Recognised procedure were applied to measure disability. Data were analysed by comprehensive statistical modelling with hierarchical structure of the data and sampling weights were considered in all analysis. Therefore, they findings are robust, and can be used in developing national level policies and programs.

However, the primary limitations of this study were the analysis of cross-sectional data, which limited our capacity to established causality and the findings were correlational only. Data were collected by asking questions to the respondents with no chance of validation. This indicates possibility of presence recall bias, though any of such bias is likely to be random. Moreover, other than the factors adjusted in the model, health and environmental factors can contribute to the onset in disability as such they are important to be adjusted in the model. However, these data were not available in the survey, limiting us to do so. However, regardless of these limitations the findings of this study will contribute to the national level policies and program development.

### Conclusions

This study reported around 3% prevalence of disability in Bangladesh with physical disability and visual impairment were the comments form of disability. Moreover, around 11% of the total disabled population reported more than one form of disability concurrently. The higher likelihoods of disability were found among comparatively higher education, higher wealth quintile and white-collar occupation holders. We also reported the divisional level variations of disability with lower likelihoods of disability were reported for the individuals in the Khulna, Rajshahi, and Rangpur divisions as well as individuals resided in the community with moderate to higher illiteracy. Awareness budling programs are important regarding the merit-based use of healthcare services rather than current tendency of need-based healthcare services use.

## Data Availability

The datasets used and analysed in this study are available at the Bangladesh Bureau of Statistics.

https://bbs.gov.bd

## Declarations

### Acknowledgement

The authors thank the Bangladesh Bureau of Statistics (BBS)for granting access to the 2021 National Survey on Persons with Disabilities data.

### Authors’ contributions

Rahman MM and Khan MN designed the study, performed the data analysis, and wrote the first draft of this manuscript. Rana MS, Rahman MM, Khandaker G and Khan MN critically reviewed and edited the previous versions of this manuscript. All authors approved this final version of the manuscript.

### Conflict of interests

The authors declare that they have no known competing financial interests or personal relationships that could have appeared to influence the work reported in this paper.

### Ethics approval and consent to participate

The survey protocol was reviewed and approved by the National Research Ethics Committee in Bangladesh.

### Consent for publication

Not applicable

Availability of data and materials: The datasets used and analysed in this study are available at the Bangladesh Bureau of Statistics.

### Conflict of 1interest

None.

## Funding

This research did not receive any specific grant from funding agencies in the public, commercial, or not-for-profit sectors.

## List of Abbreviations

PWD: Persons with disability
WHO: World Health Organization
SBI: Short Birth Interval
LMICs: Low- and middle-income countries
BBS: Bangladesh Bureau of Statistics, aOR, Adjusted Odds Ratios
AIC: Akaike Information Criteria
BIC: Bayesian Information Criteria.

## Notes

### Competing Interest Statement

The authors have declared no competing interest.

